# Tools for the assessment of quality and risk of bias in Mendelian randomization studies: a systematic review

**DOI:** 10.1101/2021.10.21.21265126

**Authors:** Francesca Spiga, Mark Gibson, Sarah Dawson, George Davey Smith, Marcus R Munafò, Julian PT Higgins

## Abstract

**Background:** The use of Mendelian randomization (MR) in epidemiology has increased considerably in recent years, with a subsequent increase in systematic reviews of MR studies. We conducted a systematic review of tools designed for risk of bias and/or quality of evidence assessment in (MR) studies, and a review of systematic reviews of MR studies.

**Methods:** We systematically searched MEDLINE, Embase, the Web of Science, preprints servers and Google Scholar for articles containing tools for assessing, conducting and/or reporting MR studies. We also searched for systematic reviews and protocols of systematic reviews of MR. From eligible articles we collected data on tool characteristics and content, as well as details of narrative description of bias assessment.

**Results:** Our searches retrieved 2464 records to screen, from which 14 tools, 35 systematic reviews and 38 protocols were included in our review. Seven tools were designed for assessing risk of bias/quality of evidence in MR studies and evaluation of their content revealed that all seven tools addressed the three core assumptions of instrumental variable analysis, violation of which can potentially introduce bias in MR analysis estimates.

**Conclusions:** We present an overview of tools and methods to assess risk of bias/quality of evidence in MR analysis. As none of these methods has been tested and validated for general use, we do not provide recommendations on their use. Our findings should raise awareness about the importance of bias related to MR analysis and provide information that is useful for assessment of MR studies in the context of systematic reviews.

## Introduction

Mendelian randomization (MR) is an analytic approach used to make causal inference in observational studies.^1^ In MR analysis, genetic variants are generally used as instrumental variables (genetic instruments, GI) to estimate the causal effect of a modifiable trait (the causal factor or “exposure”) on another trait (the factor or condition that the exposure is hypothesized to influence or “outcome”).^2^ Causal inference using MR analysis is based on the notion that genetic variants are randomly inherited from parents to offspring in a way that is comparable to participants being randomly allocated to each experimental group in a randomized controlled trial (RCT).^3^ In a within-sibship analysis randomization is almost exact,^4^ and MR was introduced through this hypothetical approach,^1^ but until recently large scale data were not available to conduct such analyses, and the approximate randomization in population-level data (adjusted for potential population stratification) has been the main approach.^3^ Thus, the key advantage of using a MR approach is the potential to reduce bias due to residual confounding and reverse causation, which are often limitations in other types of observational studies.^5^

MR was introduced as a way of strengthening causal inference regarding the kinds of modifiable exposures studied in conventional observational epidemiological studies. The key assumption here is that differences in an exposure induced by the GI will produce the same downstream effects on health outcomes as differences in the exposure produced by environmental influences (gene -environment equivalence assumption).^6^

As for instrumental variables analyses in general, the validity of an estimate from a MR analysis relies on the GI satisfying three core assumptions: (1) the GI must be associated with the exposure (*IV1-rele*vance), (2) there are no unmeasured confounders of the GI-outcome association (*IV2-independence*) and (3) the GI-outcome association must be mediated entirely via the exposure (*IV3-exclusion restriction*). Additional assumptions, which are variety of the fourth IV assumption (*IV4*),^7^ may be required for some inferences: i) the association of the GI and the exposure and the effect of the exposure on the outcome are the same for all participants in the sample (*homogeneity; ii*) the GI does not modify the effect of the exposure on the outcome within levels of the exposure and for all levels of the exposure (*no effect modification*); iii) the direction of the effect of the exposure on the outcome is the same for all participants in the sample (*monotonicity*);^8^ iiii) the differences in an exposure induced by the GI will produce the same downstream effects on health outcomes as differences in the exposure produced by environmental influences (*gene -environment equivalence assumption*).^6^ The validity of two-sample MR studies, in which different samples are used to estimate the GI-exposure and GI-outcome associations, relies on additional assumptions that the samples are independent (i.e., do not overlap); the samples are from the same underlying population (e.g., same age range) and the genetic variants are harmonised (i.e. they are in the same direction in the two samples).^9^

Violation of any of the underlying assumptions may lead to spurious or biased estimates, as may other features of the study. Some of the specific biases that have been articulated in relation to MR studies include biases emerging from the genetic instrument (e.g., weak instrument bias,^10^ bias due to horizontal pleiotropy^11^) and biases related to the population from which the data are collected (e.g., bias due to population stratification,^1,12,13^ bias due to sample overlap in two-sample MR).^14^ For example, failure to adjust for population structure and familial effects can introduce confounding in a way that is similar to lack of randomization in a RCT.^12^ Furthermore, using weak instruments in MR analysis can lead to estimates biased toward the confounded exposure-outcome association (in one-sample MR) or toward the null (in two-sample MR).^10^

Since prominent expositions of the use of MR in epidemiology from 2003 onwards,^1^ the use of MR has increased considerably, and with this has come a parallel increase in systematic reviews of MR studies. One important component of a systematic review (and meta-analysis) is the evaluation of the quality of evidence reported in each study included. This is increasingly achieved by assessing risk of bias through a structured framework. While numerous tools for risk-of-bias assessment in studies of interventions have been developed for both RCTs^15^ and non-randomized studies of intervention,^16^ and are widely used, there is no widely agreed tool for assessing MR studies.

In this systematic review we sought to identify and examine structured frameworks used to assess risk of bias (or quality more generally) in MR studies. We undertook two related sub-reviews: a comprehensive and objective review of tools for the systematic assessment of the conduct, evaluation and/or reporting of MR studies; and an examination of how risk of bias in MR studies has been assessed in systematic reviews to date.

## Methods

### Eligibility criteria

For the review of existing tools, we sought structured guidelines, checklists and other tools aimed at comprehensive evaluation of the conduct, evaluation and/or reporting of MR studies or structured guidance through the steps of conducting or reporting an MR study. For the review of systematic reviews, we examined articles describing systematic approaches to collating and summarizing MR studies within a field or more generally. We considered a systematic review any article in which the authors (i) undertook a bibliographic database search (e.g., in MEDLINE and/or other databases); and (ii) provided a table describing each of the included studies. We included full reports (e.g., full text articles) and protocols, but not conference abstracts (unless an associated full text report could be identified). We regarded any article in which genetic variants have been described or used as instrumental variables as relevant to our review.

### Searches

We performed systematic electronic searches in i) MEDLINE (Ovid), Embase (Ovid) and the Web of Science (from inception to 30 June 2021) for published peer-reviewed articles and ii) bioRxiv and medRxiv for preprint articles (last search July 2021). We implemented specific searches to identify articles describing tools (search 1), systematic reviews (search 2); and protocols for systematic reviews (search 3). To identify systematic reviews we also searched Epistemonikos, and for information on ongoing reviews we searched PROSPERO and Open Science Framework (OSF) Registries (last search 1July 2021). To identify additional articles and protocols (missed from the bibliographic database searches), we searched Google Scholar, examined references of included studies, and performed forward citation searches (Google Scholar) to identify articles citing included studies. Details of search strategies are reported in appendices 1 and 2.

### Study selection

Search results were managed using Endnote and Excel. Titles and abstracts were screened by one review author (FS) using Rayyan software (www.rayyan.ai). The full text of selected studies was retrieved and assessed for eligibility and inclusion in the review. Full text screening was performed independently by two review authors (FS and MG) and disagreements between the two reviewers were resolved through discussion. Any structured tool identified from the review of systematic reviews was incorporated into the review of tools.

### Data extraction

An extraction form was used to extract the data from the articles selected for inclusion. For each sub-review, a pilot data extraction was performed, and a finalised data extraction form was compiled. From each article, the following general information was extracted by one review author (FS): first author(s) name and year of publication, type of report (full-text article or conference abstract), type of article (e.g., tool, systematic review, protocol of systematic review) and complete reference. In addition, information specific to the two sub-reviews was extracted as follows:

#### Review of tools

number of tools within the article, purpose of the tool (i.e., conducting, evaluating, or reporting), structure of the tool (e.g., guide, dictionary, checklist), and for the evaluating tools only, specific objectives of the article, other tools used as template, number of domains and items (or questions), and specific content of each item within each tool. We extracted information only about tools designed specifically for MR studies.

#### Review of systematic reviews

review topic, whether only MR studies were included, number of included MR and non-MR studies, whether a systematic assessment of risk of bias was undertaken (or proposed if a protocol), and if applicable, whether a structured tool was used, what bias were addressed, how bias were addressed, if a narrative description of MR-specific bias was reported, and what bias where narratively addressed. We also evaluated whether a systematic assessment of the quality of evidence supporting a causal effect reported by individual MR studies was undertaken, and, if applicable, what approaches were used.

### Data analysis and reporting

We report our findings using structured summary tables and narrative descriptions. For the tools identified in the first sub-review that were aimed at the evaluation of an MR study, we tabulate the items addressed by the different tools. Where an item contained multiple questions, we separate these and tabulate each question as single item. We mapped items across tools to examine how similar bias were addressed by different tools and to convey how many of the tools addressed each bias. Specifically, we classified each item into a broad bias/topic domain, and then we assigned each item to a specific bias/topic within that domain and determined the numbers of items allocated to each bias domain and to specific MR bias/topic. For the systematic reviews identified in the second sub-review, we tabulate the methods of risk of bias and/or quality of evidence assessment in MR studies, and the MR-relevant bias addressed either by the method of assessment used or within a narrative description. For protocols of systematic reviews, we tabulate the proposed methods of assessment of risk of bias/quality of evidence in MR studies. Data extraction, narrative synthesis and tabulations were performed by one reviewer (FS).

## Results

### Tools for the conduct, evaluation and reporting of MR studies

In total, 363 records were identified from the searches (352 from database searches and 11 from other searches), of which 20 were retrieved for full-text screening. The inclusion criteria were met by 13 articles (reporting 14 tools) that are included in this review. Flow diagram of identification, screening and inclusion of articles is shown in figure 1. Of the 13 included articles, six were identified from searches of electronic databases of peer-reviewed articles and four from searches of preprints archives and Google Scholar, two from cited references, two from searches of systematic reviews (search 2) and one from searches of protocols of systematic reviews (search 3). A list of the included tools is reported in supplementary table 1.

**Figure 1:**
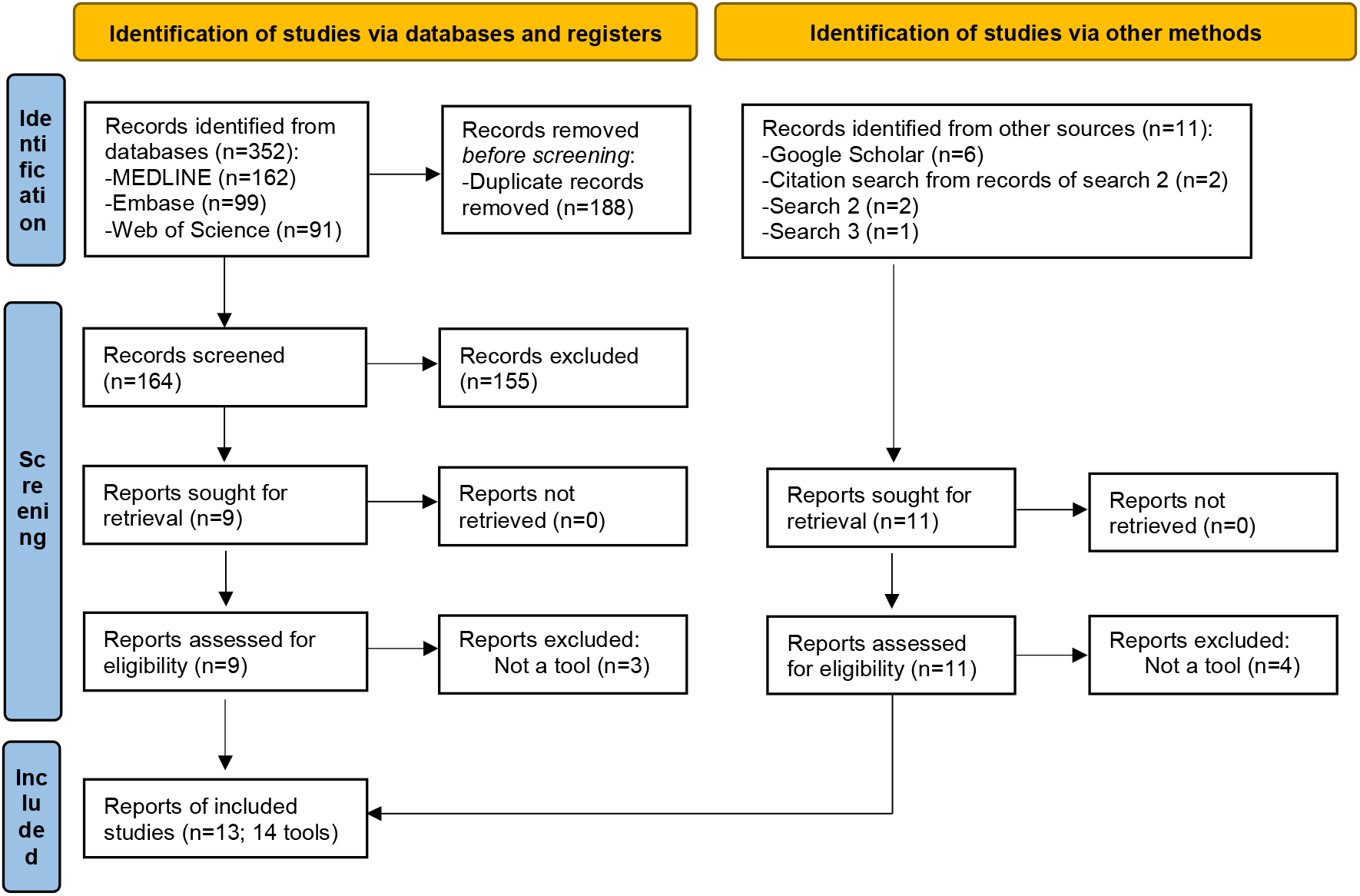
Flow diagram of identification, screening and inclusion of articles containing tools for assessing, conducting and/or reporting Mendelian randomization studies.

Of the 14 articles tools included, eight tools were designed for single use in a specific systematic review (seven reviews and one protocol) and six tools were proposed for future use for the conduct, evaluation and/or reporting of MR studies in general or within the context of a systematic review. Of the 14 identified tools, eight tools had a single purpose, of which four were aimed at the conduct of MR studies, three were aimed at the reporting of MR studies and one was aimed at evaluation of MR studies. The remaining six tools had two purposes: evaluation and reporting MR studies.

Details of the seven tools designed (or used) for evaluation of MR studies are reported in table 1. Of these, Burgess,^17^ Davies,^18^ Grau-Perez^19^ and Treur^20^ were structured by domains and items, whereas Kuźma,^21^ LS Lee^22^ and Mamluk^23^ were structured by items only. The number of domains within the first four tools ranged from 5 to 9, with a median of 6 and a total of 26 domains across the tools. The number of items in the tools ranged from 5 to 28, with a median of 19 and a total of 121 items across all the tools.

**Table 1:**
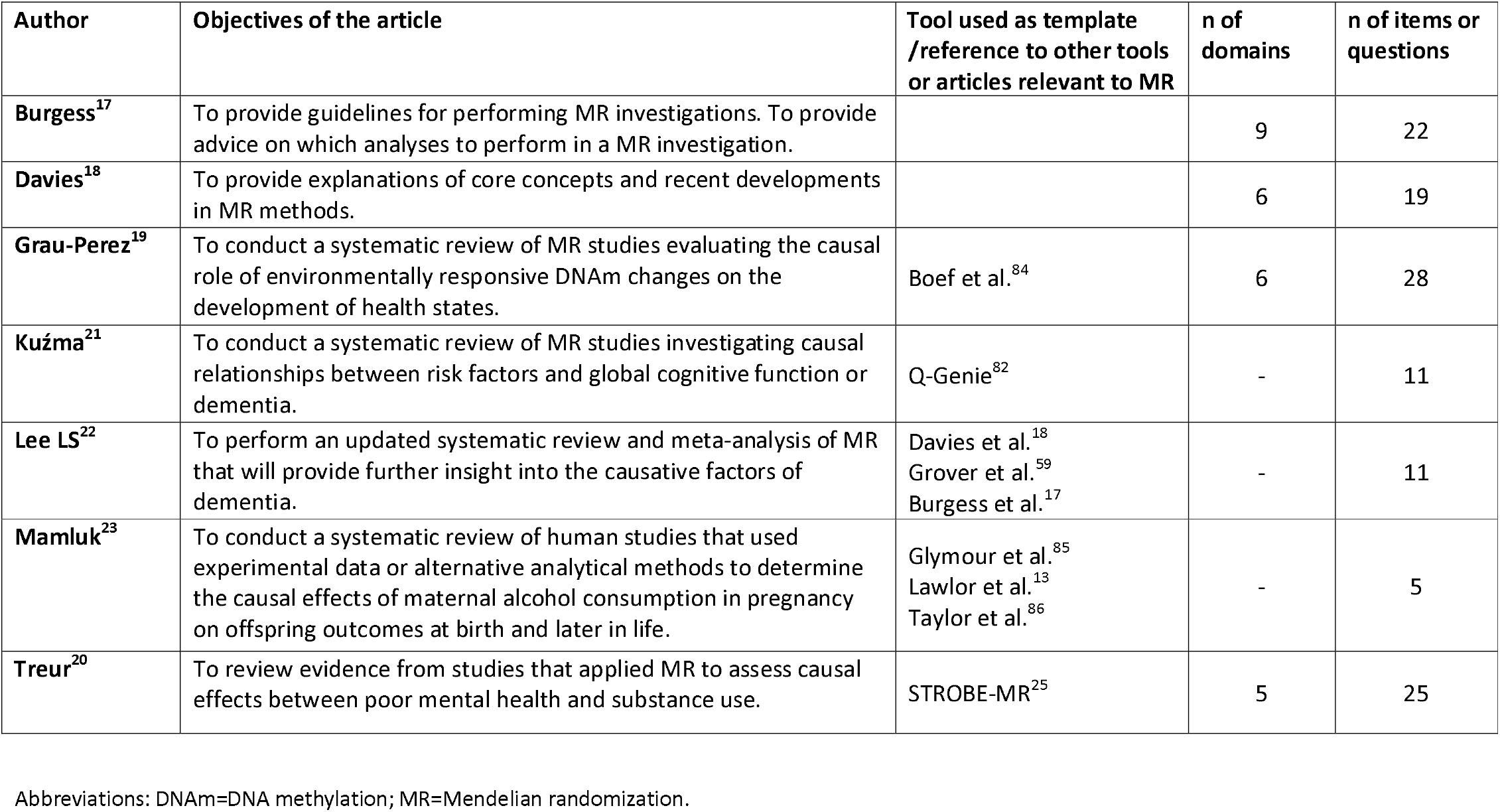
Details of tools designed/used for assessing risk of bias/evaluating quality in MR studies.

We conducted a thorough analysis of the structure and content of the evaluation tools by classifying each item into a bias/topic domain, and then we assigned each item to a specific bias/topic. We found that of the 121 items among all tools, 81 items were designed to evaluate risk of bias in MR studies, and 44 items were designed to address other aspects of the MR analysis, (four items were designed to address both evaluation of risk of bias and other aspect of MR analysis); of the 81 items designed to evaluate MR studies, 77 addressed only one bias and four addressed multiple biases.

Details of the biases addressed by each tool are reported in table 2. Of the 81 items addressing bias, 32 related to the three core IV assumptions. Ten items in seven tools addressed bias related to the relevance assumption (IV1), eight items in six tools addressed bias related to the independence assumption (IV2) and 14 items in seven tools addressed bias related to the exclusion restriction assumption (IV3). In addition, 11 items in four tools addressed bias related to the selection of the genetic instrument and 14 items in six tools addressed bias related to the selection of the population(s) or sample(s); five items in four tools addressed bias related to sensitivity analysis, 19 items in three tools addressed bias related to measurement errors and misclassification, two items in one tool addressed bias due to missing data, four items in three tools addressed bias due to other type of confounding and two items in one tool addressed other source of bias. We provide details of the 44 items addressing other aspect of the MR analysis, including items addressing the reporting of MR analysis, in <u>supplementary table 2</u>. Among these, we found that two items in one tool addressed clinical implications of the MR results; three items in three tools addressed the choice of dataset(s); four items in three tools addressed the genetic instrument; six items in two tools addressed the interpretation of the MR analysis results; five items in three tools addressed the MR rationale; six items in three tools addressed the MR results; four items in three tools addressed precision of the results; two items in one tool addressed the selection of the population(s) or sample(s) and seven items in four tools addressed the statistical analysis.

**Table 2:** Details specific MR bias and limitation addressed by items or questions within each assessing tool.

| Bias (or topic) domain | Specific bias (or topic) | Burgess | Davies | Grau-Perez | Kuzma | Lee | Mamluk | Treur | Total items |
| --- | --- | --- | --- | --- | --- | --- | --- | --- | --- |
| **IV1-Relevance** | Choice of variants | Y |  |  |  |  |  |  | 1 |
|  | Weak instrument bias |  | Y | Y | Y | Y | Y | Y | 9 |
| **IV2-Independence** | Choice of variants | Y |  |  |  |  |  |  | 1 |
|  | Confounding |  | Y | Y | Y | Y<sup>b</sup> | Y<sup>c</sup> | Y | 7 |
|  | Population stratification |  |  |  |  |  | Y |  | 1 |
| **IV3-Exclusion restriction** | Choice of variants | Y |  |  |  |  |  |  | 1 |
|  | Horizontal pleiotropy | Y | Y | Y | Y | Y | Y | Y | 13 |
| **Genetic instrument** | Choice of variants | Y |  |  |  |  |  |  | 3 |
|  | Construction of genetic score |  |  |  |  |  |  | Y | 1 |
|  | Variants harmonization | Y | Y |  |  | Y |  | Y | 4 |
|  | Linkage disequilibrium | Y | Y |  |  | Y |  |  | 3 |
| **Population/sample** | Samples overlap<sup>a</sup> | Y | Y |  |  | Y |  | Y | 5 |
|  | Population heterogeneity<sup>a</sup> | Y | Y | Y |  | Y | Y | Y | 6 |
|  | Choice of controls |  |  | Y |  |  |  |  | 1 |
|  | Selection bias |  |  | Y |  |  |  |  | 1 |
| **Sensitivity analysis** | Evidence of robustness | Y | Y |  |  | Y |  | Y | 5 |
| **Measurement error** | Exposure measurement error/misclassification |  |  | Y | Y |  |  | Y | 13 |
|  | Outcome measurement error/misclassification |  |  | Y | Y |  |  | Y | 6 |
| **Missing data** | - |  |  | Y |  |  |  |  | 2 |
| **Other confounding** | - |  |  | Y | Y |  | Y |  | 4 |
| **Other sources of bias** | - |  |  |  | Y |  |  |  | 2 |

<sup>a</sup>In two-sample MR analysis. <sup>b</sup>Confounding of the genetic instrument-outcome association. <sup>c</sup>Confounding of the genetic instrument-exposure association and of the genetic instrument-outcome association. Abbreviations: IV1=instrumental variable assumption 1; IV2=instrumental variable assumption 2; IV3=instrumental variable assumption 3.

In addition to the evaluation tools, we identified three tools aimed at reporting and four tools aimed at conducting MR studies; all seven tools contained items addressing bias in MR analysis and details of the content of the items is reported in <u>supplementary table 3</u>. The number of domains ranged from three to six in the reporting tools and from five to ten in the conducting tools; the number of items ranged from seven to 61 in the reporting tools and from 18 to 26 in the conducting tools. Among the reporting tools, all three tools contained items addressing the three IV core assumptions, Boef^24^ contained items addressing linkage disequilibrium and canalization; Davey Smith^25^ contained items addressing homogeneity and sample overlap (in two-sample MR); Lor^26^ contained items addressing linkage disequilibrium and heteroscedasticity. Among the conducting tools, Burgess,^17^ Grover^27^ and Lawlor^28^ contained items addressing the three IV core assumptions, and variant harmonization; in addition, Burgess^17^ contained one items addressing the homogeneity assumptions and Grover^27^ and Lawlor^28^ contained items addressing sample overlap; Swerdlow^29^ contained items addressing linkage disequilibrium and horizontal pleiotropy.

### Systematic reviews of MR studies

#### Completed reviews

A total of 2036 record were identified from searches 2 (for systematic reviews) (2025 from database searches and 11 from other searches) of which 143 were retrieved for full-text screening, and the inclusion criteria were met by 38 articles (35 full-text articles, and 3 conference abstracts linked to included articles) reporting 35 reviews that are included in this synthesis. A flow diagram of identification, screening and inclusion of studies is shown in figure 2. A list of included reviews is reported in table 3. Of the 35 included reviews, 25 were systematic reviews and ten were umbrella reviews. Of the 35 included reviews, 29 addressed a clinical question (i.e., included studies on the casual effect of an exposure vs an outcome), and six reviews addressed a methodological question (e.g., the status of reporting in MR studies); 17 reviews reported MR studies only, the other 18 reported both MR and non-MR studies; the number of MR studies ranged between 1 and 231 with a median of 18 studies. Of the 35 included reviews, 14 conducted an assessment of either risk of bias or quality of the evidence: six reviews conducted risk-of-bias assessments only, five reviews conducted quality of evidence assessments only and three did both. Details of the risk of bias and quality of evidence assessment in individual MR studies used in these 14 reviews are reported in <u>supplementary table 4.</u>

**Figure 2:**
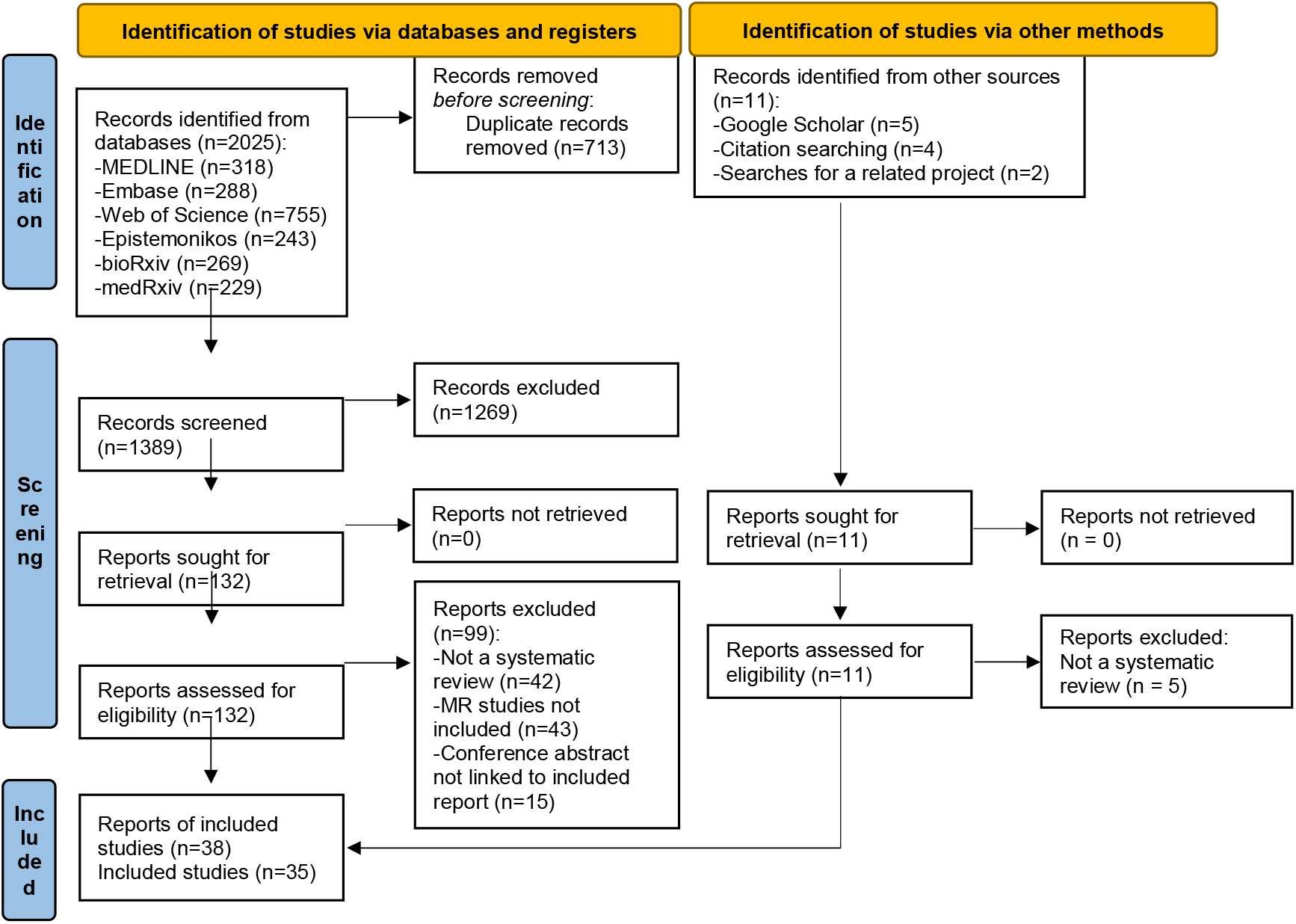
Flow diagram of identification, screening and inclusion of articles containing systematic reviews (and meta-analysis) of Mendelian randomization studies

**Table 3:** List of included systematic reviews reporting one or more Mendelian randomization studies.

| Study ID | Type of article | Topic of the review | Were only MR studies included? | N of MR studies/N of non-MR studies | Risk of bias assessment in individual MR studies? If Yes, was a structured tool used? | Name and/or description of risk of bias assessment method | Evidence of causal effect assessment in individual MR studies? If Yes, was a structured method used? | Description of evidence of causal effect assessment method | Narrative description of MR-specific bias |
| --- | --- | --- | --- | --- | --- | --- | --- | --- | --- |
| Abbasi<sup>87</sup> | Systematic review and MR analysis | MR studies of biomarkers and T2D | Yes | 28/0 | No | N/A | No | N/A | Yes |
| Abbasi<sup>88</sup> | Systematic review | Biomarkers and T2D | No | 17/122 | No | N/A | No | N/A | Yes |
| Belbasi<sup>89</sup> | Umbrella review | Risk factors and peripheral biomarkers for schizophrenia and other psychotic disorders | No | 5/36 | No | N/A | No | N/A | No |
| Belbasi<sup>90</sup> | Umbrella review | Risk factors of multiple sclerosis | No | 6/9 | No | N/A | No | N/A | Yes |
| Bellou<sup>91</sup> | Umbrella review | Environmental risk factors and biomarkers for T2D | No | 22/86 | No | N/A | No | N/A | Yes |
| Bergmans<sup>92</sup> | Systematic review | Comorbid depression and T2D | No | 4/12 | No | N/A | No | N/A | Yes |
| Bochud<sup>37</sup> | Literature review on MR methods, applications, and limitations | MR studies | Yes | 38/0 | No | N/A | Yes | Strength of genetic variant | Yes |
| Boef<sup>84</sup> | Systematic review | Methodology used in MR analysis | Yes | 179/0 | No | N/A | No | N/A | Yes |
| Carnegie<sup>93</sup> | Literature review on MR methods, applications and limitations and systematic review of MR studies | MR in Nutritional psychiatry | Yes | 26/0 | No | N/A | No | N/A | Yes |
| Cheng<sup>30</sup> | Systematic review and meta-analysis | Puberty timing and T2D and/or impaired glucose tolerance | No | 1/27 | Yes, Yes | Newcastle-Ottawa Scale<sup>31</sup> | No | N/A | No |
| Diemer<sup>94</sup> | Systematic review | Prenatal environment and offspring outcomes | Yes | 43/0 | No | N/A | No | N/A | Yes |
| Firth<sup>38</sup> | Umbrella review | Modifiable health behaviors and major mental disorders | No | 12/32 | No | N/A | Yes | Statistical analysis results, use of sensitivity analysis and | Yes |

|  |  |  |  |  |  |  |  | test for<br>bidirectional<br>effects |  |
| --- | --- | --- | --- | --- | --- | --- | --- | --- | --- |
| Frayling<sup>95</sup> | Systematic<br>review | MR studies of<br>T2D, coronary<br>artery disease<br>and hypertension | Yes | 16/0 | No | N/A | No | N/A | Yes |
| Grau-Perez<sup>a19</sup> | Systematic<br>review | MR studies of<br>environmentally<br>responsive<br>DNAm changes<br>and the<br>development of<br>health states | Yes | 15/0 | Yes, Yes | Self-developed<br>tool | No | N/A | Yes |
| Hu<sup>96</sup> | Systematic<br>review | MR studies of<br>atherosclerotic<br>cardiovascular<br>disease | Yes | 58/0 | No | N/A | No | N/A | Yes |
| Kei<sup>97</sup> | Systematic<br>review | MR studies of<br>serum uric acid<br>levels and<br>cardiovascular<br>and renal disease<br>risk | Yes | 16/0 | No | N/A | No | N/A | Yes |
| Kim<sup>39</sup> | Umbrella<br>review of<br>systematic<br>reviews and<br>meta-analyses | Adiposity and<br>cardiovascular<br>disease events or<br>mortality | No | 27/11 | No | N/A | Yes | Statistical<br>power | Yes |
| Kohler<sup>40</sup> | Umbrella<br>review of<br>meta-analysis<br>and MR | Environmental<br>risk factors for<br>depression | No | 8/70 | No | N/A | Yes | Proportion<br>of variance<br>in risk<br>factors | No |

|  | studies |  |  |  |  |  |  | explained by genetic instruments |  |
| --- | --- | --- | --- | --- | --- | --- | --- | --- | --- |
| Kuzma<sup>a21,98</sup> | Systematic review | MR studies of risk factors and global cognitive function or dementia | Yes | 18/0 | Yes, Yes | Modified Q-Genie<sup>82</sup> | No | N/A | Yes |
| Li<sup>41</sup> | Umbrella review of systematic reviews and meta-analyses | Serum uric acid level and multiple health outcomes | No | 36/101 | No | N/A | Yes | Statistical significance of the effect estimate and statistical power | Yes |
| Lor<sup>a26</sup> | Systematic review | MR analyses in oncological studies | Yes | 77/0 | No | N/A | No | N/A | Yes |
| Mamluk<sup>a23,99</sup> | Systematic review | Maternal alcohol consumption in pregnancy and offspring outcomes at birth and later in life | No | 9/14 | Yes, Yes | Self-developed tool | No | N/A | Yes |
| Markozannes<sup>32</sup> | Umbrella review | C-reactive protein and health outcomes | No | 37/55 | Yes, No | Assessment of horizontal pleiotropy<sup>b</sup> | Yes/Yes | Statistical significance of the effect estimate | Yes |
| Meng<sup>100</sup> | Systematic review of MR studies and MR analysis | MR studies of vitamin D and health outcomes | Yes | 65/0 | No | N/A | No | N/A | No |
| Pearson-Stuttard<sup>34</sup> | Umbrella review | T2D and cancer incidence or mortality | No | 8/20 | Yes, No | Assessment of selection of genetic instrument | Yes/Yes | Statistical significance of the effect estimate | Yes |
| Pingault<sup>101</sup> | Systematic review | MR studies of psychopathology-related outcomes | Yes | 19/0 | No | N/A | No | N/A | Yes |
| Riaz<sup>35,36</sup> | Systematic review and meta-analysis of MR studies | MR studies of obesity and CVD | Yes | 7/0 | Yes, No | Evaluation of the three MR core assumptions | No | N/A | Yes |
| Robinson<sup>102</sup> | Literature review on MR methods, applications and limitations and systematic review of MR studies | MR studies of rheumatology | Yes | 33/0 | No | N/A | No | N/A | Yes |
| Sommer<sup>103</sup> | Systematic review | Childhood and adolescent obesity and future cardiovascular morbidity and mortality later in life | No | 1/85 | No | N/A | No | N/A | No |
| Swerdlow<sup>a29</sup> | Review on methods for selecting instruments for MR analysis and Systematic review of MR studies | MR studies | Yes | 231/0 | No | N/A | No | N/A | Yes |
| Treur<sup>a20</sup> | Systematic review | MR studies of poor mental health and substance use | Yes | 63/0 | Yes, Yes | Self-developed tool | No | N/A | Yes |
| Vasta<sup>104</sup> | Systematic review | Diabetes mellitus and amyotrophic lateral sclerosis | No | 1/35 | No | N/A | No | N/A | No |
| Yuan<sup>105</sup> | Systematic review and MR analysis | MR studies of risk factors of T2D | Yes | 40/0 | No | N/A | No | N/A | No |
| Zhang X<sup>106</sup> | Umbrella review | Non-genetic biomarkers and colorectal cancer | No | 18/78 | Yes, No | Assessment of horizontal pleiotropy | Yes/Yes | Statistical significance of the effect estimate, statistical power and evidence of bias due to directional pleiotropy | Yes |
| Zhang Z<sup>33</sup> | Systematic review | Vitamin D and non-alcoholic fatty liver disease | No | 1/12 | No | N/A | No | N/A | No |

<sup>a</sup>Included in the synthesis of tools for the assessing/evaluating MR studies. <sup>b</sup>Based on the location of the SNPs. Abbreviations: DNAm=DNA methylation; MR=Mendelian randomization; N/A=not applicable; SNP=single nucleotide polymorphism; T2D=type 2 diabetes.

A structured risk-of-bias tool for was used in five reviews: four of these (Grau-Perez,^19^ Kuzma,^21^ Mamluk^23^ and Treur^20^) used tools developed specifically for risk-of-bias assessment in MR studies that are included in the above sub-review of tools (see supplementary rable 1 and table 2); the fifth, Cheng,^30^ used the Newcastle Ottawa Scale (NOS) for cohort studies^31^ which was not specifically developed for MR studies. Four further reviews conducted risk-of-bias assessments but did not use a structured tool: Markozannes^32^ and X Zhang^33^ assessed horizontal pleiotropy; Pearson-Stuttard^34^ addressed the selection of the genetic instrument(s); and Riaz^35,36^ conducted evaluation of the three core assumptions.

Of the eight reviews that conducted a quality of evidence assessment, Markozannes^32^ and Pearson-Stuttard^34^ used a structured method based on statistical significance of the effect estimate and X Zhang^33^ used a structured method based on a combination of statistical significance of the effect estimate, statistical power and evidence of bias due to directional pleiotropy. Among the other five reviews in which a structured method was not used, Bochud^37^ based the assessment of quality of evidence on the strength of the genetic variant; Firth^38^ based the assessment on the results of the statistical analysis, the use of sensitivity analysis and test for bidirectional effects; Kim^39^ based the assessment on statistical power; Kohler^40^ based the assessment on the proportion of variance in risk factors explained by genetic instruments used and Li^41^ based the assessment on the statistical significance of the effect estimate and the statistical power.

Of the 35 reviews included, 28 reported a general narrative description of potential bias and limitation in MR studies. Details of specific biases addressed narratively within these systematic reviews are reported in <u>supplementary table 3</u>. Of these 28 reviews, 20 addressed bias related to the IV1 assumption (i.e., weak instrument bias), 16 reviews addressed bias related to the IV2 assumption (i.e., confounding, population stratification, assortative mating, dynastic effect and parent of origin effect),^12^ and 24 reviews addressed bias related to the IV3 assumption (i.e., horizontal pleiotropy). In addition, 17 reviews addressed bias related to the selection of the genetic instrument (i.e., linkage disequilibrium, Winner’s course bias, segregation distortion, monotonicity and homogeneity), six reviews addressed bias related to the selection of the population or sample (i.e., population heterogeneity and selection bias), eight reviews addressed bias due to canalization, and four reviews addressed bias due to measurement errors or misclassification. In addition to bias, we also evaluated whether other MR-relevant topics were narratively described, and we found that 11 reviews addressed precision of the results (i.e., low statistical power or sample size), five reviews addressed reverse causation (or bidirectionality), three reviews addressed the inability to assess non-linear associations, two reviews addressed statistical analysis and lack of genetic instrument, respectively, and one review addressed inability to assess dose-response estimations.

#### Protocols for systematic reviews

Our final search for protocols of systematic reviews (search 3) identified 65 protocols (57 from database searches and 8 from other searches, including 1 from search 2) of which 15 were excluded because inclusion of MR studies was not specified, or MR studies were specified in the exclusion criteria. A flow diagram of identification, screening and inclusion of protocols of systematic reviews is shown in figure 3. Two protocols for the same review were identified from different sources for five reviews, therefore a total of 45 study protocols were included in this part of the review. A list of included protocols with details of the method used by each of study is reported in table 4. Five of the 45 included protocols were of published systematic review that were included in our sub-review of systematic reviews above.^42-46^ Of the 45 included protocols, 35 were for systematic reviews of primary studies and 10 were for umbrella reviews. Fifteen protocols were for reviews of MR studies only and 30 planned to include other study designs.

**Figure 3:**
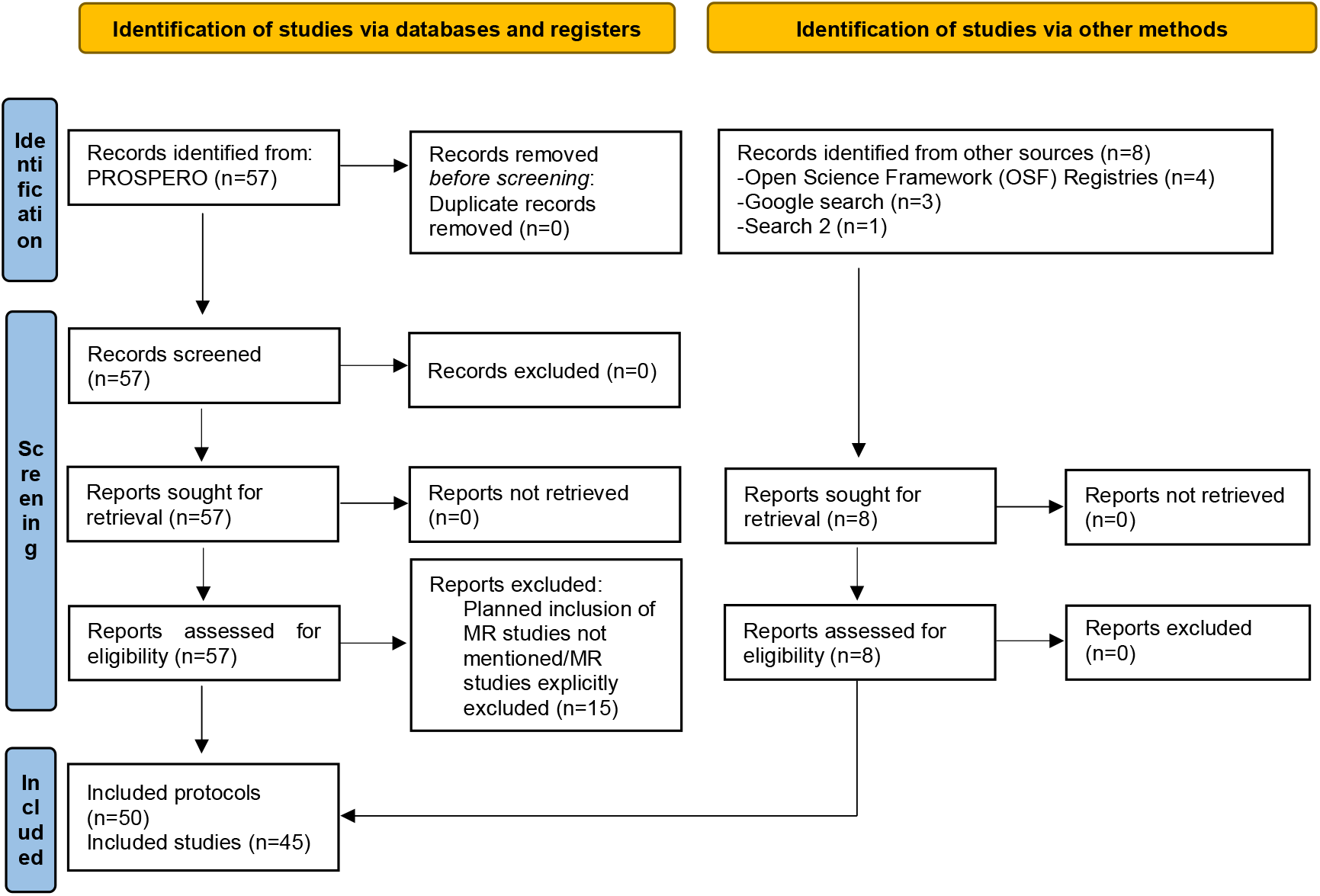
Flow diagram of identification, screening and inclusion of protocols of systematic reviews (and meta-analysis) planning to include Mendelian randomization studies.

**Table 4:**
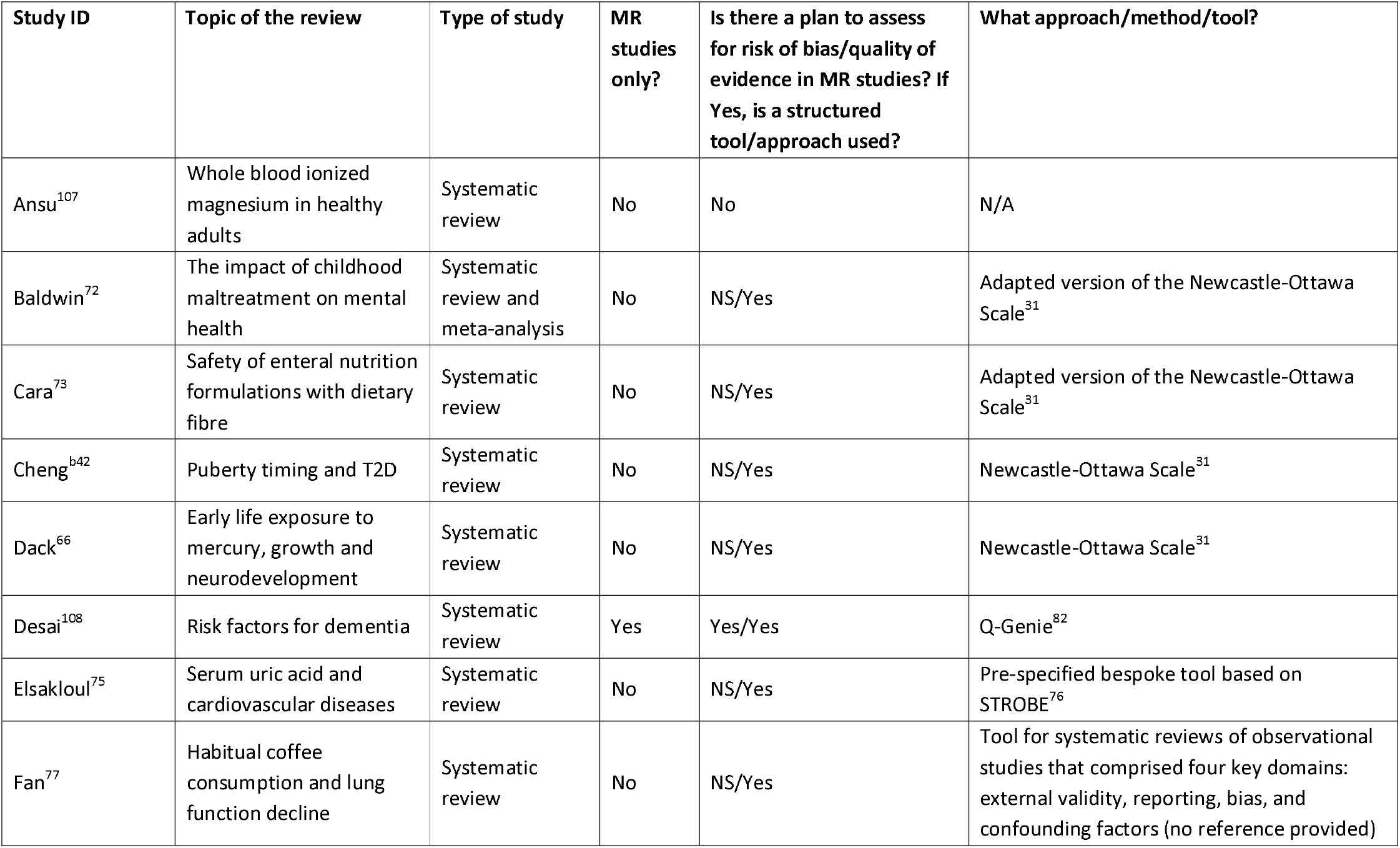
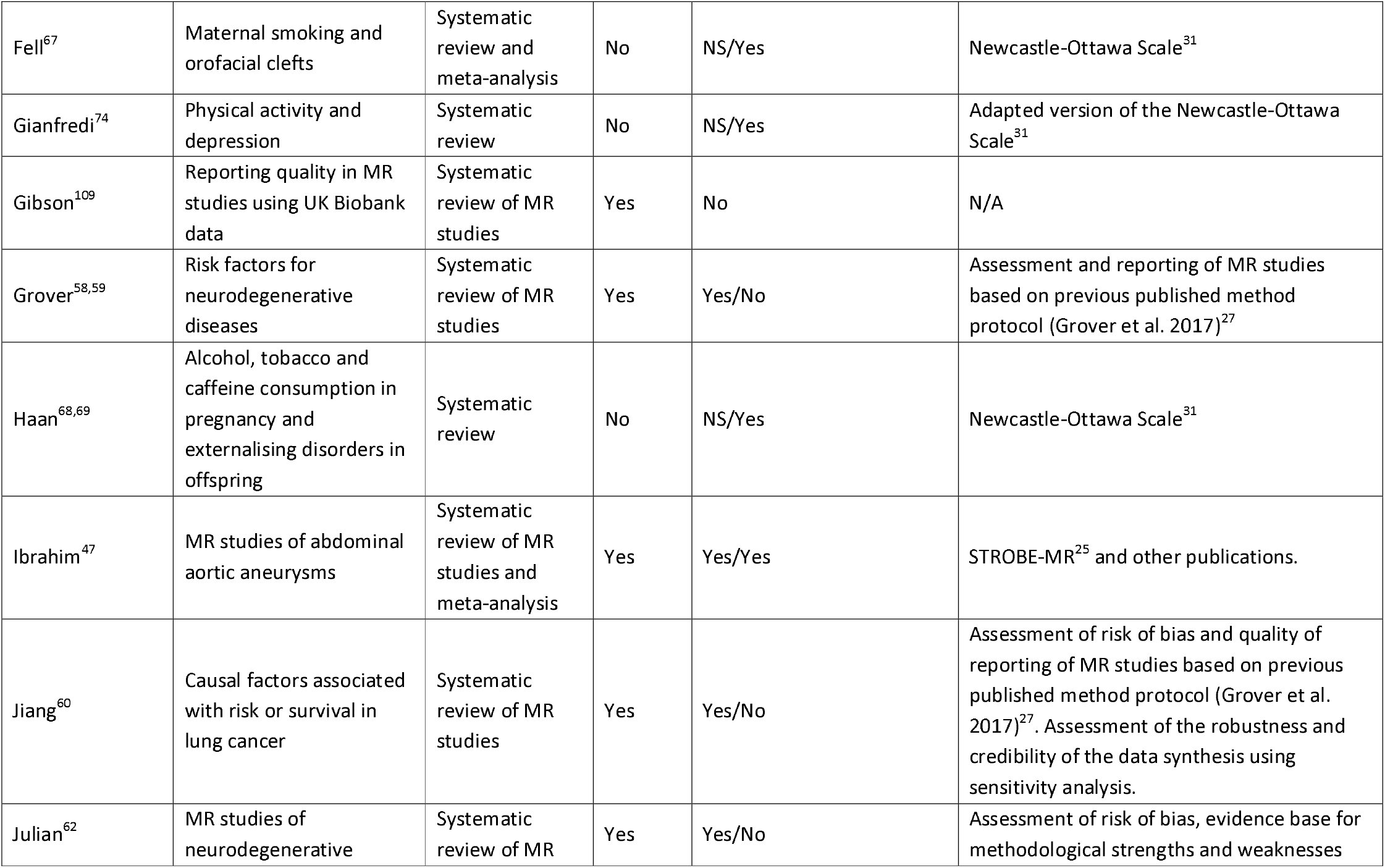
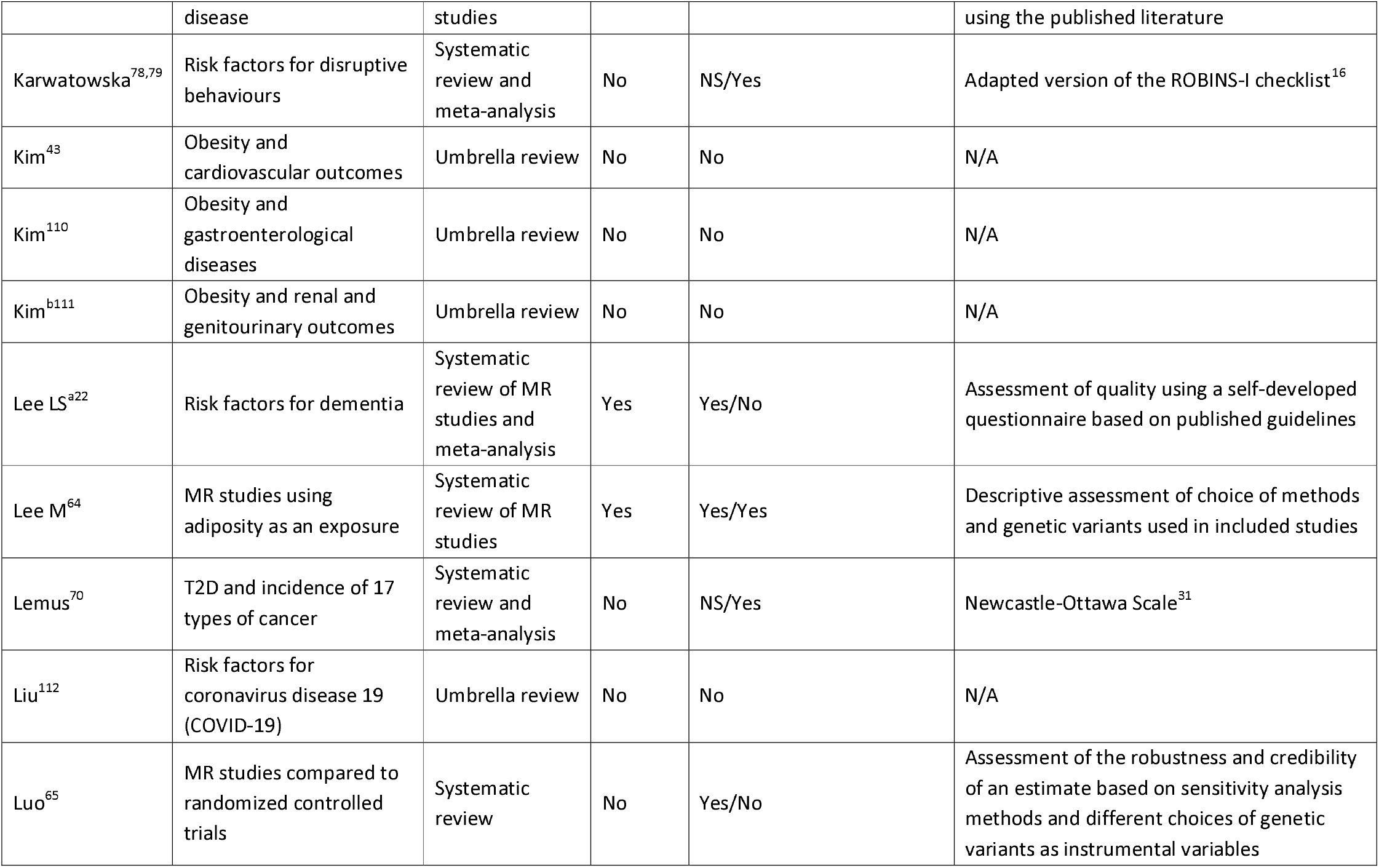
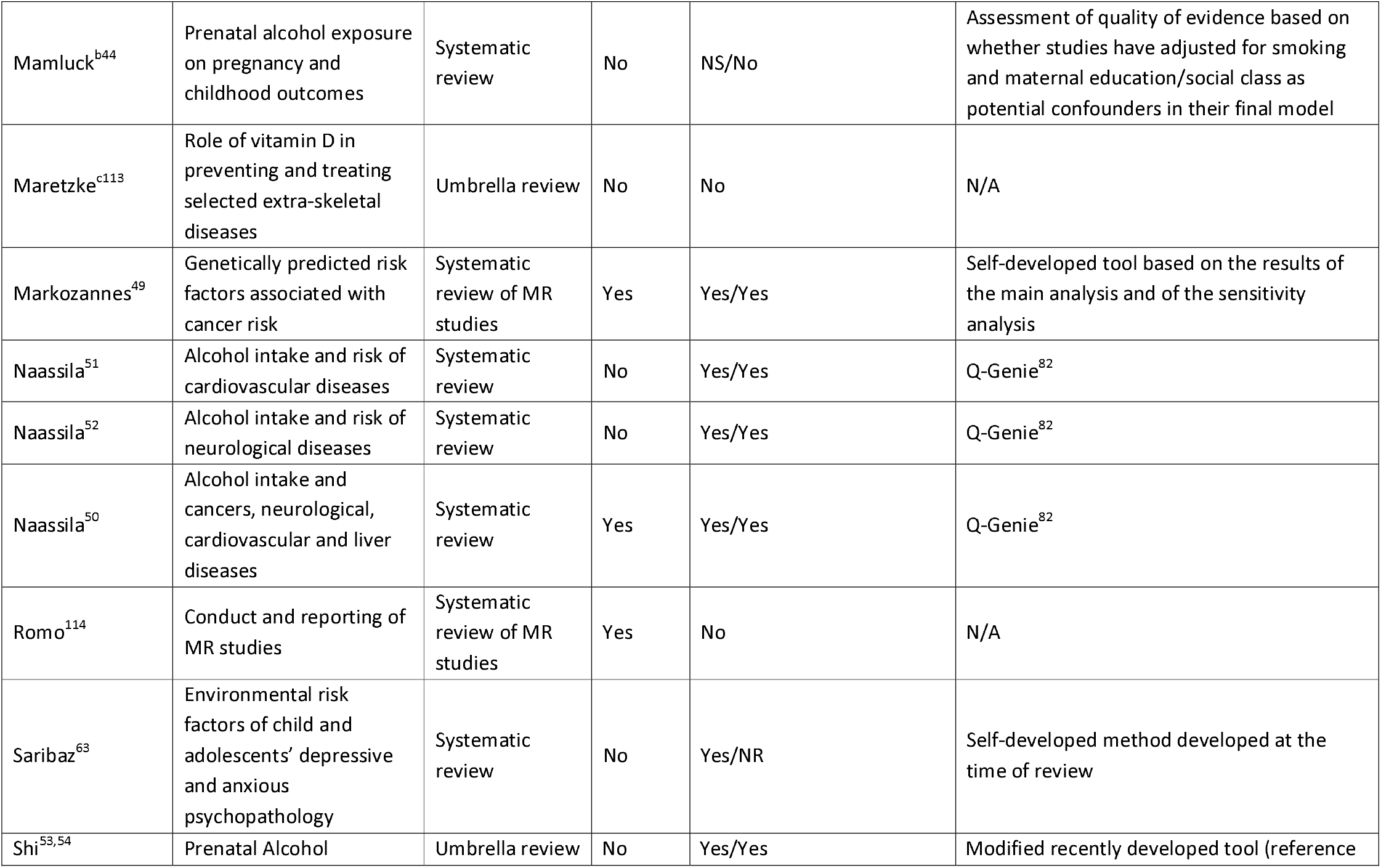
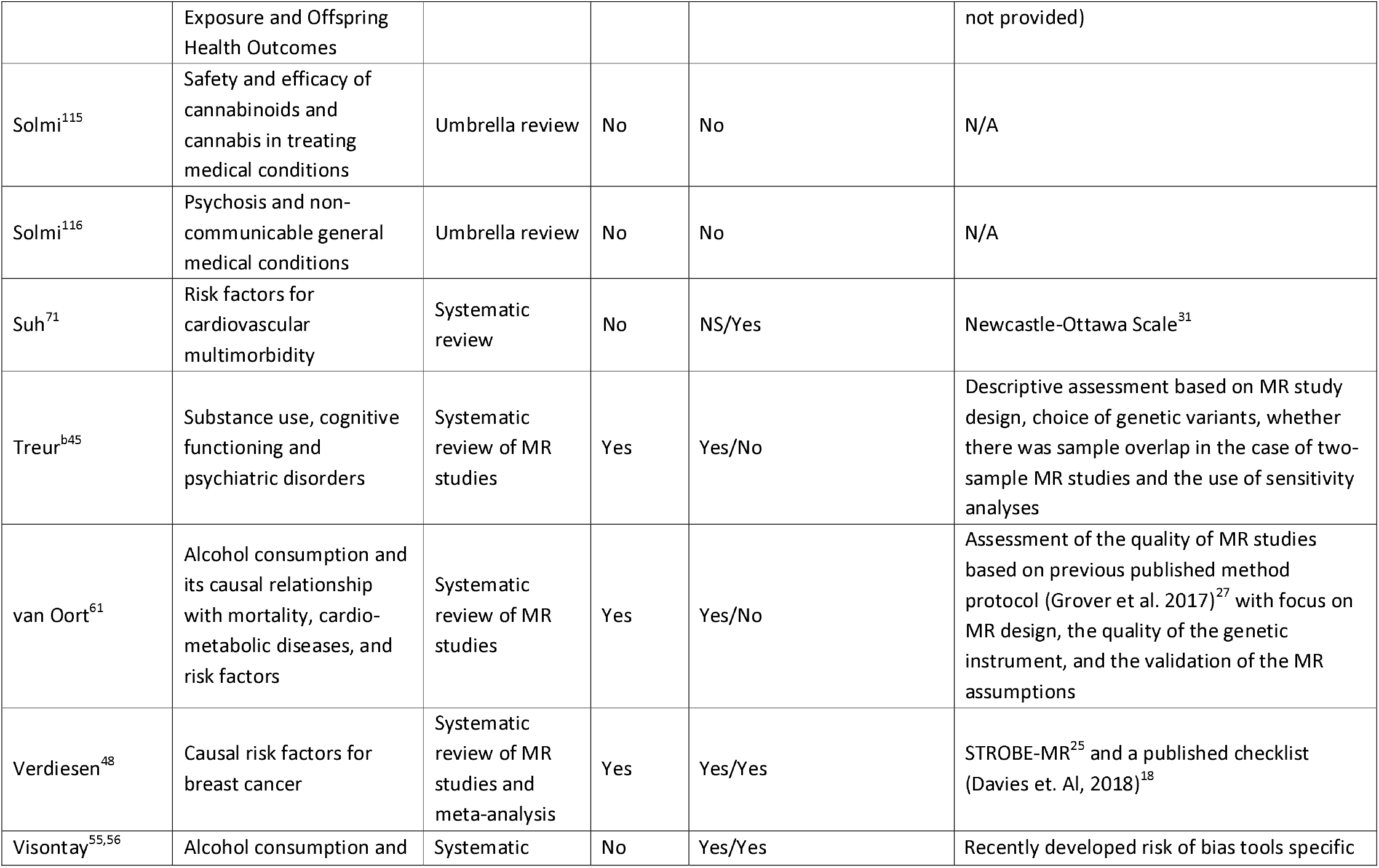
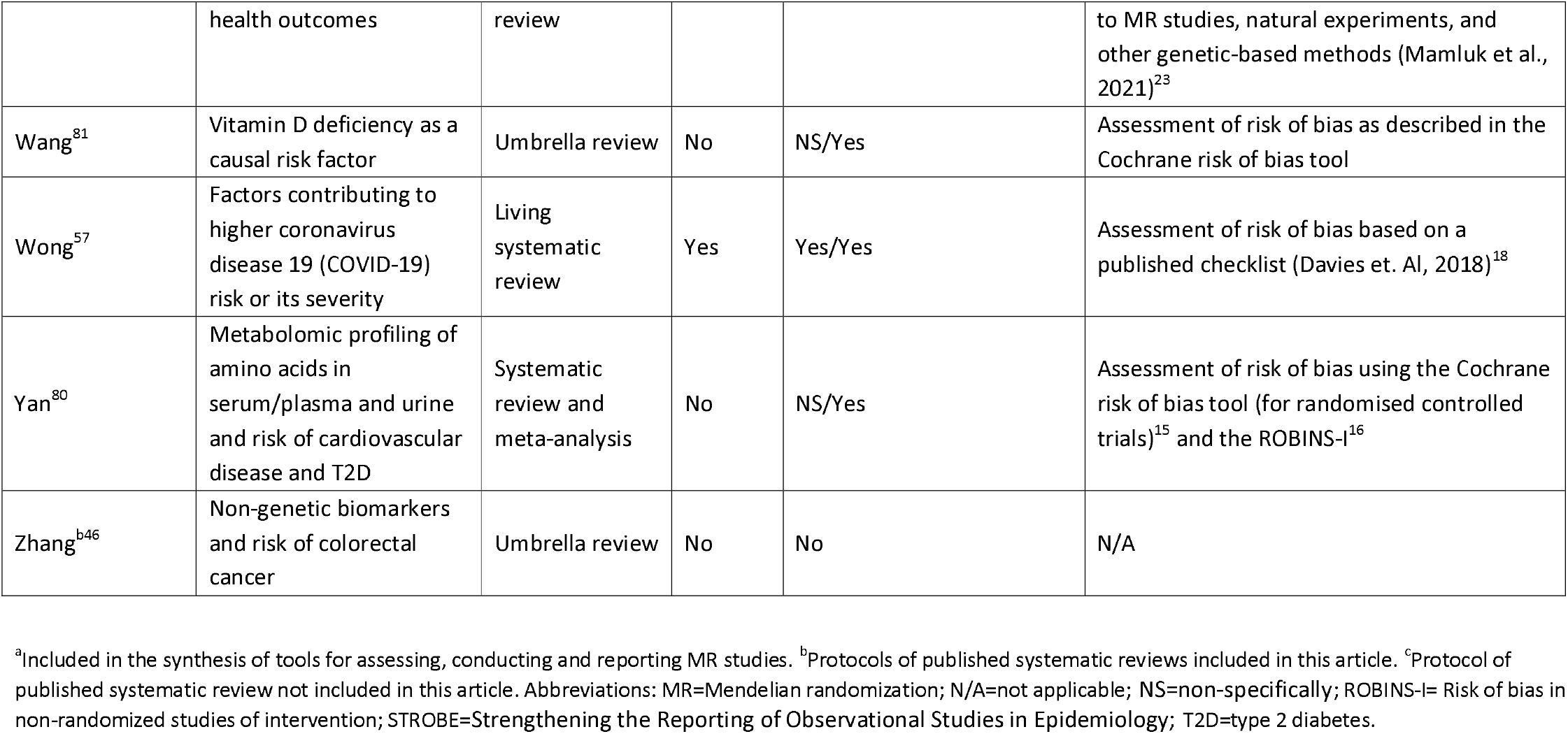
List of included protocols of systematic reviews reporting MR studies.

Eighteen protocols reported plans for a MR-specific risk-of-bias/quality-of-evidence assessment and 15 protocols reported plans for a non-MR-specific risk-of-bias/quality-of-evidence assessment. Of the 18 protocols with a MR-specific risk-of-bias/quality-of-evidence assessment, the use of a structured tool/method was planned in 11 protocols, the use of other methods/approaches was planned in 12 protocols and one protocol described the use of a method that the author planned to develop at the time of conducting the review. Of the 11 protocols describing use of a structured tool, Ibrahim^47^ and Verdiesen^48^ planned to use STROBE-MR^25^ and other published literature, including the MR guidelines by Davies,^18^ LS Lee^22^ planned to use a self-developed questionnaire (also included in our synthesis of tools) based on published guidelines including Davies,^18^ Grover,^27^ and Burgess.^17^ Markozannes^49^ planned to use a self-developed tool based on the results of the main analysis and of the sensitivity analysis; Naassila^50-52^ planned to use Q-GENIE; Shi^53,54^ planned to use a modified version of a recently developed tool (no reference provided); Visontay^55,56^ planned to use the tool developed by Mamluk^23^ and Wong^57^ planned to conduct risk of bias assessment based on the guidelines from Davies.^18^ Of the seven protocols describing a MR-specific risk-of-bias/quality-of-evidence assessment without using a structured tool, four planned an assessment based on the literature: Grover, ^58,59^ Jiang^60^ and van Oort^61^ referred to the MR methods protocol published by Grover,^27^ and Julian^62^ did not report any reference. Of the remaining four protocols, Saribaz^63^ planned to develop a risk-of-bias assessment method at the time of conducting the review; M Lee^64^ planned to perform a descriptive assessment of the MR methods and of the genetic variants used in included studies; Luo^65^ planned to perform an assessment based on sensitivity analysis methods and different choices of genetic variants as instrumental variables; Treur^45^ planned to perform an assessment based on sensitivity analysis methods, on the choice of genetic variants, on the presence of sample overlap (two-sample MR studies) and on the use of sensitivity analyses.

Of the 15 protocols in which a non-MR-specific risk-of-bias assessment is reported, 14 used structural tools and Mamluk^44^ planned to assess risk of bias on whether adjustment for potentially relevant confounders was conducted. Of the 14 structured tools used for non-MR-specific risk-of-bias assessment, Cheng,^42^ Dack,^66^ Fell,^67^ Haan,^68,69^ Lemus^70^ and Suh^71^ planned to use NOS,^31^ and Baldwin,^72^ Cara^73^ and Gianfredi^74^ planned to use a modified version of NOS; Elsakloul^75^ planned to use STROBE,^76^ Fan^77^ planned to use a quality-assessment tool for systematic reviews of observational studies that comprised external validity, reporting, bias, and confounding factors, but a reference was not provided; Karwatowska^78,79^ planned to use ROBINS-I,^16^ Yan^80^ planned to use the ROB-2^15^ and the ROBINS-I^16^ tools, Wang^81^ planned to use the Cochrane risk-of-bias assessment tool (no details provided).

## Discussion

Our systematic review of tools developed for the conduct, evaluation and/or reporting of MR studies identified 14 instruments. Half of the tools were designed (or used) either entirely or partially for the evaluation of MR studies. Most of these tools were developed for application within a systematic review,^19-23^ whereas only two were developed for general use.^17,18^ Despite notable variability in the structure and content of the tools, all tools contained items addressing the validity of the three core IV assumptions. In addition, all but one of the tools addressed bias related to the selection of the population(s) or sample(s), including population heterogeneity, sample overlap, choice of controls and selection bias, and just over half of the tools addressed bias related to the genetic instrument, including linkage disequilibrium, construct of the genetic score and lack of variants harmonization, and addressed the conduct of sensitivity analysis. Fewer than half of the tools addressed bias due to measurement errors and only one tool addressed bias due to other sources including missing data. While it was not in our scope to critically appraise the identified tools, by compiling a list and inspecting the content of these tools we found that all tools, including these designed for reporting and conducting, addressed these assumptions or conditions within the MR analysis that, when violated, lead to potential bias of the MR causal estimate.

Consistent with the lack of formal tools for assessment of risk of bias in MR studies, only a small proportion (26%) of the systematic reviews of MR studies included in our review conducted a risk-of-bias assessment, and only 23% of the included reviews conducted an assessment of evidence of causal effect within individual MR studies. Nevertheless, most of the reviews included a narrative description of MR-related bias and limitations (74%), and – as observed in the content of the tools – among these, most of the reviews addressed bias related to the core IV assumptions of relevance (IV1) and exclusion restriction (IV3) (71% and 86% respectively), but only 57% addressed bias related to the independence assumption (IV2), whereas 61% addressed bias related to the genetic instrument and only 21% addressed bias related to the selection of the population or sample.

In contrast with published systematic reviews, when we looked at protocols of systematic reviews of (or including) MR studies, a plan to conduct an assessment was reported in 73% of the protocols included in our reviews, although only in 40% the approach or methodology used was specific for MR studies. This higher proportion may reflect an increased focus on risk of bias over time or may reflect a tendency for review teams who publish their protocols to include risk-of-bias assessments in their plans. Of protocols that specified methodologies specific to MR studies, only 39 % planned to use a structured tool, including the STROBE-MR,^25^ Q-GENIE,^82^ a self-developed tool included in our synthesis of tools^22^ and a tool developed within another systematic review.^23^ One review protocol planned to use a recently developed tool, that, similarly to the tool developed by Mamluk,^23^ consisted of five questions, one for bias domain, including instrument bias, genetic confounding, and selection bias. The rest of the protocols not planning to use a structured tool proposed other informal ways to address bias, including assessment based on the validation of the three IV core assumptions, the choice of genetic instruments, the use of sensitivity analysis and description of MR analysis design, and some of these approaches were based on MR literature including MR guidelines by Davies^18^ and Grover.^27^

Our review has strengths and limitation. First, we included published and unpublished articles by searching several relevant databases for peer-reviewed articles, preprints archives and Google Scholar for preprints articles and unpublished studies. Furthermore, for each objective, specific search string developed with the assistance of an information specialist. However, as some of the tools we have identified were developed within other type of articles, including literature reviews and systematic reviews of MR and non-MR studies, it is possible that our searches may have missed some tools. As data extraction was performed by a single author, it is possible that some errors in data collection were made. Our classification of items into bias domains and specific issues is to an extent arbitrary, and some items could have been classified in accordance with more than one bias or limitation. For example, although we classified linkage disequilibrium as relevant to the choice of genetic variant, it can introduce both confounding^5^ and horizontal pleiotropy^83^ and therefore it can be considered as bias related to the IV2 or IV3 domains.

By summarising the currently available knowledge on methods and approaches for assessment of risk of bias in MR studies, our longer-term aim was to identify potential items for inclusion in a structured tool for risk-of-bias assessment in MR studies. Given that none of the tools identified by our searches appears to have been formally tested, we are not able to make a recommendation on what tool(s) should be adopted to assess MR studies. However, the content of the tools that we have identified in our review will be a useful source of information on what bias/limitations reviewers should be aware of when conducting a systematic review (and meta-analysis) including results from MR studies, and what types of biases reviewers should consider when assessing the quality of the evidence reported by individual MR studies.

## Data Availability

All data produced in the present study are available upon reasonable request to the authors

