## Appendix for "Tools for the assessment of quality and risk of bias in Mendelian randomization studies: a systematic review"

**Appendix 1. Search strategy for tools aimed at evaluation of the design, conduct and/or reporting of Mendelian randomization studies (search 1)**

**Ovid MEDLINE ALL and Embase**

1. Mendelian randomi#ation.hw,tw,kw,kf.

2. genetic instrument?.hw,tw,kw,kf.

3. (genetic variant* adj3 instrumental variable*).tw,kw,kf.

4. or/1-3

5. scale*. tw,kw,kf.

6. checklist*. tw,kw,kf.

7. critical apprais*.tw,kw,kf.

8. tool*. tw,kw,kf.

9. guide*. tw,kw,kf.

10. (Dictionary or glossary). tw,kw,kf.

11. or/5-10

12. valid*. hw,tw,kw,kf.

13. quality. hw,tw,kw,kf.

14. ((bias OR confound*) AND (asses* OR measur* OR evaluat*)).tw,kw,kf.

15. or/12-14

16. 4 and 11 and 15

**Fields: *hw*** ([subject] heading word); ***kf*** (keyword heading word (MEDLINE)); ***kw*** (keyword (Embase); keyword heading (MEDLINE)); **tw** (text word).

**Web of Science**

(**TI**=((("Mendelian randomization" or "Mendelian randomisation" or "genetic instrument*" or ("genetic variant*" and "instrumental variable*")) AND (scale* or checklist* or “critical apprais*” or tool* or guide* or dictionary or glossary) AND (valid* or quality or ((bias or confound*) and (asses* or measur* or evaluat*)))))) OR

(**AB**=((("Mendelian randomization" or "Mendelian randomisation" or "genetic instrument*" or ("genetic variant*" and "instrumental variable*")) AND (scale* or checklist* or “critical apprais*” or tool* or guide* or dictionary or glossary) AND (valid* or quality or ((bias or confound*) and (asses* or measur* or evaluat*)))))) OR (**AK**=((("Mendelian randomization" or "Mendelian randomisation" or "genetic instrument*" or ("genetic variant*" and "instrumental variable*")) AND (scale* or checklist* or “critical apprais*” or tool* or guide* or dictionary or glossary) AND (valid* or quality or ((bias or confound*) and (asses* or measur* or evaluat*))))))

**Fields: TI**: title; **AK**: author keyword; **AB**: abstract.

**Appendix 2. Search strategy for systematic reviews of Mendelian randomization studies (search 2)**

**Ovid MEDLINE ALL and Embase**

1. Mendelian randomi#ation.hw,tw,kw,kf.

2. genetic instrument?.hw,tw,kw,kf.

3. (genetic variant* adj3 instrumental variable*).tw,kw,kf.

4. or/1-3

5. ((systematic or meta) adj2 (analys* or review)).ti,kw,kf.

6. ((systematic* or quantitativ* or methodologic*) adj5 (review* or overview*)).ti,ab,kw,kf,sh.

7. (quantitativ$ adj5 synthesis$).ti,ab,kw,kf,hw.

8. meta-analysis.pt.

9. (meta-analys* or meta analys* or metaanalys* or meta synth* or meta-synth* or metasynth*).ti,ab,kw,kf,hw.

10. ((umbrella or overview) adj3 (review? or meta-analys*)).mp.

"review of meta-analyses".mp.

11. (2046-4053).is. [ISSN for Systematic Reviews journal]

12. or/ 5-13

13. 4 and 14

**Fields: *hw*** ([subject] heading word); ***kf*** (keyword heading word (MEDLINE)); ***kw*** (keyword (Embase); keyword heading (MEDLINE)); ***tw*** (text word); ***ti*** (title); ***ab*** (abstract); ***mp*** (multiple places search – which also includes ***ot*** (original title); ***ox*** (other index terms word (Embase))); ***is*** (ISSN prints (MEDLINE); ISSN (Embase)).

**Web of Science**
**(TI**=(("Mendelian randomization" or "Mendelian randomisation" or "genetic instrument*" or ("genetic variant*" and "instrumental variable*")) AND (("systematic or meta" NEAR "analys* or review") or (systematic* or quantitativ* or methodologic* NEAR review* or overview*) or (meta-analys* or meta analys* or metaanalys* or meta synth* or meta-synth* or metasynth*)))) OR (**AK**=(("Mendelian randomization" or "Mendelian randomisation" or "genetic instrument*" or ("genetic variant*" and "instrumental variable*") ) AND (("systematic or meta" NEAR "analys* or review") or (systematic* or quantitativ* or methodologic* NEAR review* or overview*) or (meta-analys* or meta analys* or metaanalys* or meta synth* or meta-synth* or metasynth*)))) OR (**AB**=(("Mendelian randomization" or "Mendelian randomisation" or "genetic instrument*" or ("genetic variant*" and "instrumental variable*") ) AND (("systematic or meta" NEAR "analys* or review") or (systematic* or quantitativ* or methodologic* NEAR review* or overview*) or (meta-analys* or meta analys* or metaanalys* or meta synth* or meta-synth* or metasynth*))))

**Fields: TI**: title; **AK**: author keyword; **AB**: abstract.

**bioRxiv and medRxiv:** Mendelian AND randomi*ation AND review

**Search strategy for protocols of systematic reviews of Mendelian randomization studies (search 3):**

**PROSPERO**: Mendelian randomization OR Mendelian randomisation.
