## Supplementary tables 1-4 for "Tools for the assessment of quality and risk of bias in Mendelian randomization studies: a systematic review"

**Supplementary table 1: List of included studies containing one or more tools for evaluating, conducting, and reporting MR studies**

| **Study ID** | **Type of article** | **Scope of the tool(s)** | **Number of tools** | **Structure of the tool** |
| --- | --- | --- | --- | --- |
| **Boef^a1^** | Systematic review of methodological approaches and quality of reporting in MR studies | Reporting | 1 | Checklist |
| **Burgess^2^** | Guideline for performing MR investigations | Assessing  Conducting  Reporting | 2 | Domain-based checklist (assessing and reporting tool)  Flowchart (conducting tool) |
| **Davey Smith^3^** | STROBE-MR; guideline for the reporting of MR studies | Reporting | 1 | Checklist |
| **Davies^4^** | Guide, checklist and glossary for MR studies | Assessing  Reporting | 1 | Checklist |
| **Grau-Perez^a5^** | Systematic review of MR studies | Assessing  Reporting | 1 | Domain-based evaluation chart |
| **Grover^6^** | Guideline for performing MR analysis | Conducting | 1 | Flowchart |
| **Kuźma^a7^** | Systematic review of MR studies | Assessing  Reporting | 1 | Rating scale |
| **Lawlor^8^** | MR dictionary and online tool | Conducting | 1 | Flowchart |
| **Lee^b9^** | Protocol for a systematic review of MR studies on risk factors for dementia | Assessing  Reporting | 1 | Questionnaire |
| **Lor^a10^** | Systematic review of MR studies and guideline for reporting MR studies | Reporting | 1 | Checklist |
| **Mamluk^a11^** | Systematic review of MR studies | Assessing | 1 | Bias domain-based rating |
| **Swerdlow^a12^** | Review of selection of instrument in MR analysis and systematic review of MR studies | Conducting | 1 | Decision tree |
| **Treur^a13^** | Systematic review of MR studies | Assessing  Reporting | 1 | Scoring system |

^a^ Included in the synthesis of systematic review of MR studies. ^b^ Included in the synthesis of protocols of systematic reviews of MR studies. Abbreviations: MR=Mendelian randomization.

**Supplementary table 2: Details of other MR-relevant content of items or questions within each assessing tool**

| **Aspect of MR analysis** | **Specific topic** | **Burgess** | **Davies** | **Grau-Perez** | **Kuzma** | **Lee** | **Mamluk** | **Treur** | **Total** |
| --- | --- | --- | --- | --- | --- | --- | --- | --- | --- |
| **Clinical implications** | Reporting of clinical implication |  | Y |  |  |  |  |  | 2 |
| **Datasets** | Reporting of datasets used |  | Y |  |  | Y |  |  | 2 |
| **Genetic instrument** | Reporting of MR rationale (biological rationale) | Y |  |  |  |  |  |  | 1 |
|  | Reporting of method used to obtain genetic variant |  |  | Y |  |  |  |  | 1 |
|  | Reporting of genetic variant strength |  |  |  |  |  |  | Y | 2 |
| **Interpretation** | Applicability/transportability |  | Y |  |  |  |  |  | 1 |
|  | Interpretation of results |  | Y |  |  |  |  |  | 2 |
|  | Reporting of MR estimates for interpretation of results |  | Y |  |  |  |  |  | 1 |
|  | Bidirectional effects |  |  |  |  |  |  | Y | 1 |
|  | Temporality |  |  |  |  |  |  | Y | 1 |
| **MR rationale** | Reporting of MR rationale | Y |  |  | Y |  |  | Y | 5 |
| **MR results** | Reporting of comparison of MR estimate with observational study estimate |  | Y | Y |  | Y |  |  | 3 |
|  | Reporting of MR analysis results |  | Y | Y |  |  |  |  | 3 |
| **Precision** | Statistical power/sample size |  |  | Y | Y |  |  | Y | 4 |
| **Selection of population/sample** | Gathering specific methods details | Y |  |  |  |  |  |  | 1 |
|  | Reporting of relevance of the selected population to the research question | Y |  |  |  |  |  |  | 1 |
| **Statistical analysis** | Reporting of statistical methods |  |  | Y | Y |  |  |  | 2 |
|  | Reporting of MR analysis results | Y |  |  |  |  |  |  | 1 |
|  | Reporting of primary analysis statistical methods | Y |  |  |  |  |  |  | 1 |
|  | Reporting of primary analysis details | Y |  |  |  |  |  | Y | 2 |
|  | Reporting of secondary analysis statistical methods | Y |  |  |  |  |  |  | 1 |
| **Type of dataset** | Gathering specific methods details | Y |  |  |  |  |  |  | 1 |

Abbreviations: MR=Mendelian randomization; Y=yes.

**Supplementary table 3: Details of reporting and conducting tools**

| **Author** | **Objectives of the tool** | **Scope of the tool** | **Tool used as template /reference to other tools or articles relevant to MR** | **n of domains** | **n of items** | **Details of bias addressed** |
| --- | --- | --- | --- | --- | --- | --- |
| **Boef** | To provide an overview of the use of the different approaches to MR and the specific statistical methods used for estimation. To evaluate whether the plausibility of the MR assumptions is discussed. To evaluate whether the statistical methods used are sufficiently described. | Reporting | NR | 3 | 7 | Weak instrument; confounders of GI-exposure association; unmeasured confounders of GI-outcome association; population stratification; horizontal pleiotropy; linkage disequilibrium, canalization; |
| **Davey Smith (STROBE-MR)** | To provide guidelines for strengthening the reporting of MR studies. | Reporting | STROBE ^14^ | 6 | 44 | IV1-3; homogeneity; sample overlap; |
| **Lor** | To evaluate the reporting quality of MR analyses in cancer studies and provide guidelines for reporting in MR biomedical research, for use in future publications. | Reporting | PRISMA ^15^  STREGA ^16^  STROBE ^14^ | 6 | 44^a^; 61^b^ | Weak instrument; confounders of GI-exposure and/or GI-outcome association; population stratification; horizontal pleiotropy; linkage disequilibrium; heteroscedasticity; |
| **Burgess** | To provide guidelines for performing MR investigations. To provide advice on which analyses to perform in a MR investigation. | Conducting | NR | 9 | 26 | IV1-3 assumptions; variants harmonization; homogeneity assumption; |
| **Grover** | To provide a step-by-step guide for causal inference based on the principles of MR with a real dataset using both individual and summary data from unrelated individuals. | Conducting | NR | 6 | 24 | Sample overlap; variants harmonization; confounders of GI-exposure and/or GI-outcome association; strength of genetic variant; horizontal pleiotropy; |
| **Lawlor** | To provide definitions and key references for terms and concepts that are necessary to undertake MR studies, and to critically appraise and appropriately interpret findings from MR studies. | Conducting | NR | 10 | 18 | Sample overlap; variants harmonization; confounders of GI-exposure and/or GI-outcome association; strength of genetic variant; horizontal pleiotropy; |
| **Swerdlow** | To provide guidance that may help investigators to plan a Mendelian randomization study of a disease-associated biomarker, and readers interpret Mendelian randomization studies. | Conducting | NR | 5 | 20 | Linkage disequilibrium; horizontal pleiotropy; |

Abbreviations: GI=genetic instrument; IV=instrumental variable; MR=Mendelian randomization; NR=not reported; PRISMA=Preferred Reporting Items for Systematic Reviews and Meta-Analyses; STREGA= STrengthening the REporting of Genetic Association studies; STROBE=Strengthening the Reporting of Observational Studies in Epidemiology. **^a^**Studies with data pooling; ^b^Studies without data pooling.

**Supplementary table 4: Details of bias addressed by a narrative description in systematic reviews of MR studies.**

|  | **IV1 (Relevance)** | **IV2**  **(Independence)** | | | | | **IV3**  **(Exclusion restriction)** |
| --- | --- | --- | --- | --- | --- | --- | --- |
| **Study ID** | **Weak instrument** | **Confounding** | **Population stratification** | **Assortative mating** | **Dynastic effect** | **Parent of origin effect** | **Horizontal pleiotropy** |
| Abbasi |  | Y |  |  |  |  | Y |
| Abbasi |  |  |  |  |  |  | Y |
| Belbasi | Y | Y |  |  |  |  | Y |
| Bellou | Y |  |  |  |  |  |  |
| Bergmans |  |  |  |  |  |  | Y |
| Bochud | Y | Y | Y |  |  | Y | Y |
| Boef | Y | Y | Y |  |  |  | Y |
| Carnegie | Y |  | Y |  |  |  | Y |
| Diemer | Y | Y | Y | Y |  |  | Y |
| Firth |  |  |  |  |  |  | Y |
| Frayling | Y | Y | Y |  |  |  | Y |
| Grau-Perez | Y | Y | Y |  |  |  | Y |
| Hu | Y |  | Y |  |  |  | Y |
| Kei | Y |  |  |  |  |  | Y |
| Kim | Y |  |  |  |  |  |  |
| Kohler |  |  |  |  |  |  |  |
| Kuzma | Y |  | Y |  |  |  | Y |
| Li | Y | Y |  |  |  |  | Y |
| Lor | Y | Y^a^ | Y | Y |  |  | Y |
| Mamluk | Y | Y | Y | Y | Y |  | Y |
| Markozannes |  |  |  |  |  |  | Y |
| Pearson-Stuttard |  |  |  |  |  |  | Y |
| Pingault | Y |  | Y |  |  |  | Y |
| Riaz | Y |  |  |  |  |  | Y |
| Robinson | Y | Y | Y |  |  |  | Y |
| Swerdlow | Y |  |  |  |  |  | Y |
| Treur |  | Y | Y | Y | Y |  | Y |
| Zhang | Y |  |  |  |  |  |  |

**Supplementary table 4 (continued)**

|  | **Selection of genetic instrument** | | | | | **Selection of sample** | |
| --- | --- | --- | --- | --- | --- | --- | --- |
| **Study ID** | **Linkage disequilibrium** | **Winner's curse^b^** | **Segregation distortion** | **Lack of monotonicity** | **Allelic homogeneity** | **Population heterogeneity** | **Selection bias** |
| Abbasi |  |  |  |  |  |  |  |
| Abbasi |  |  |  |  |  |  |  |
| Belbasi |  |  |  |  |  |  |  |
| Bellou |  |  |  |  |  |  |  |
| Bergmans |  |  |  |  |  |  |  |
| Bochud | Y |  | Y |  | Y |  | Y^c^ |
| Boef |  | Y |  |  |  |  |  |
| Carnegie | Y |  |  |  |  |  |  |
| Diemer |  | Y |  |  |  | Y | Y |
| Firth |  |  |  |  |  |  |  |
| Frayling |  |  |  |  |  |  |  |
| Grau-Perez | Y |  |  |  |  |  |  |
| Hu | Y |  |  |  |  |  |  |
| Kei | Y |  |  |  |  |  |  |
| Kim |  |  |  |  |  |  |  |
| Kohler |  |  |  |  |  |  |  |
| Kuzma |  |  |  |  |  | Y | Y |
| Li |  |  |  |  |  |  |  |
| Lor | Y |  |  |  |  | Y |  |
| Mamluk |  |  |  |  |  |  | Y |
| Markozannes |  |  |  |  |  |  |  |
| Pearson-Stuttard |  |  |  | Y |  |  |  |
| Pingault | Y |  |  |  |  |  |  |
| Riaz |  |  |  |  |  |  |  |
| Robinson | Y |  |  |  |  |  |  |
| Swerdlow | Y | Y |  |  |  |  |  |
| Treur | Y |  |  |  |  |  | Y |
| Zhang | Y |  |  |  |  |  |  |

**Supplementary table 4 (continued)**

|  | **Canalization** | **Measurement errors / misclassification** | **Lack of genetic instrument** | **Reverse causation (or bidirectionality)** | **Inability to assess non-linear associations** | **Inability to assess dose–response estimation** | **Low statistical power** | **Statistical analysis** |
| --- | --- | --- | --- | --- | --- | --- | --- | --- |
| Abbasi | Y |  |  |  |  |  |  |  |
| Abbasi | Y |  |  |  |  |  |  |  |
| Belbasi |  |  |  |  |  |  |  |  |
| Bellou |  |  |  |  |  |  | Y |  |
| Bergmans |  |  |  |  |  |  |  |  |
| Bochud | Y |  |  |  |  |  |  |  |
| Boef |  |  |  |  |  |  |  |  |
| Carnegie | Y |  | Y |  | Y |  | Y |  |
| Diemer |  | Y |  |  |  |  | Y | Y |
| Firth |  |  |  | Y | Y | Y | Y |  |
| Frayling |  |  |  |  |  |  |  |  |
| Grau-Perez |  |  |  |  |  |  |  |  |
| Hu | Y |  |  |  |  |  | Y |  |
| Kei | Y |  |  |  |  |  |  |  |
| Kim |  |  |  | Y |  |  | Y |  |
| Kohler |  |  |  |  |  |  | Y |  |
| Kuzma |  |  |  |  |  |  |  |  |
| Li |  |  |  |  |  |  |  |  |
| Lor |  |  |  |  |  |  | Y |  |
| Mamluk | Y | Y |  | Y | Y |  | Y |  |
| Markozannes |  |  |  |  |  |  |  |  |
| Pearson-Stuttard |  | Y |  |  |  |  |  |  |
| Pingault |  | Y |  | Y |  |  | Y |  |
| Riaz |  |  |  |  |  |  |  |  |
| Robinson | Y |  |  |  |  |  | Y |  |
| Swerdlow |  |  |  |  |  |  |  |  |
| Treur |  |  |  | Y |  |  |  |  |
| Zhang |  |  |  |  |  |  |  | Y |

^a^ Confounding by shared genetics and environment. ^b^ In selecting genetic instruments**;** ^c^ Selective survival due to the genetic variant of interest. Abbreviations: IV1= instrumental variable assumption 1; IV2= instrumental variable assumption 2; IV3= instrumental variable assumption 3; Y=yes.

9.	Lee LS, Whiteley W, Walker R. Systematic review and meta-analysis of Mendelian randomisation studies on modifiable risk factors for dementia. protocols.io

<https://dx.doi.org/10.17504/protocols.io.bpeemjbe>.
